# Comparison of COVID-19 Home-Testers vs Laboratory-Testers in New York State (Excluding New York City), November 2021 to April 2022

**DOI:** 10.1101/2022.10.21.22281319

**Authors:** Vajeera Dorabawila, Virgile Barnes, Nirmala Ramesh, Rebecca Hoen, Jamie Sommer, Amy Robbins, Byron Backenson, Emily Lutterloh, Dina Hoefer, Eli Rosenberg

**Affiliations:** New York State Department of Health, ESP Corning Tower Rm 754, Albany, NY 12237

## Abstract

**Background:** Though the use of home testing is increasing, it is not represented in the reported coronavirus disease 2019 (COVID-19) metrics. As the epidemic and its tracking evolve, it is critical to understand who the excluded home-tested persons are relative to those in reported metrics.

**Methods:** Five New York State databases were linked to understand the trends in home-tested COVID-19 cases compared to laboratory-confirmed cases from November 2021 to April 2022. Frequency distributions, logistic regression adjusted odds ratios (aOR), and 95% confidence intervals (CI) were used to compare the characteristics of home-tested and laboratory-tested persons.

**Results:** Of 592,227 confirmed COVID-19 cases, 71,531 (12%) had a home-test-only, 515,001 (87%) had a laboratory-test-only and 5,695 (1%) had both a home-test and laboratory-test during this period. Home-tested cases, as a percentage of confirmed COVID-19 cases, increased from 1% in November 2021 to 22% in April 2022. Children aged 5-11 years with 3.74 aOR (95% CI:3.53, 3.96) and adolescents aged 12-17 years with 3.24 aOR (95% CI:3.07, 3.43) were more likely to have home-test-only than adults aged 65 and above. Boosted (aOR 1.87, 95% CI:1.82, 1.93), in K-12 school settings (aOR 2.33, 95% CI:2.27, 2.40), or possibly infected by a household member (aOR 1.17, 95% CI:1.13, 1.22) were more likely to report home-test-only than laboratory-test-only. Individuals hospitalized (aOR 0.04, 95% CI:0.03, 0.06), with underlying conditions (aOR 0.85, 95% CI:0.83, 0.87), pregnant (aOR 0.76, 95% CI:0.66, 0.86), Hispanic (aOR 0.50:95% CI:0.48, 0.53), Asian (aOR 0.31, 95% CI:0.28, 0.34), or Black (aOR 0.45, 95% CI:0.42, 047) were less likely to utilize the home test only compared to the laboratory test only.

**Conclusion:** The number of individuals using home tests only as a proportion of confirmed COVID-19 cases continues to increase. Home test-only cases are less likely to be hospitalized and have a lower potential for severe disease as measured by age, vaccination status, and underlying conditions. Thus, those with severe disease and the potential for severe disease are represented as official metrics. Racial and ethnic differences exist between persons reporting home and laboratory tests.

## 1. Introduction

The coronavirus disease 2019 (COVID-19) epidemic has persisted, and there is a continued need to monitor its trends. As of September 21, 2022, there were almost 6 million COVID-19 infections and 400,000 reinfections in the NYS. At-home rapid COVID-19 antigen test (home-test) availability and usage have increased rapidly in the United States with the emergence of the B.1.2.529 (Omicron) variant in late 2021 (1-8). As case volume surged with Omicron in late 2021 and January 2022, a shortage of laboratory-based diagnostic nucleic acid amplification (laboratory-based tests) testing relative to demand resulted in increased home test usage (6). A recent national study reported that at-home test usage by those with COVID-19-like illness increased from approximately 5.7% during the delta dominant period (August 23, 2021– December 11, 2021) to 20.1% in the Omicron dominant period (December 19, 2021–March 12, 2022) (9). Another study reported an increasing trend in home-test use and found that during the week ending on January 8, 2022, when testing volume peaked, the positivity rate was 17.3 for home tests and 29.1 for laboratory-based tests (10). Home tests are easily accessible, offer privacy during testing, and provide rapid results compared to laboratory-based tests. Simultaneously, an increase in accessibility was driven by the school, state, and federal government-free test kits, and increased volume in pharmacies and retail venues (11).

Official reporting, as well as other analytics, are primarily based on laboratory-based testing, as home tests are not typically reported to data systems tracking COVID-19 case volume and rates. Consequently, there is a dearth of information on home test usage and users; studies available are very few, survey-based with an associated response bias and are on adults (12, 13). Given this scenario, there is a critical need to better understand the individuals, including children, using and reporting home tests, as well as trends in home tests. Doing so will shed light on the potential bias, if any, in analyses based solely on laboratory testing.

This paper explores several questions associated with home tests in NYS, excluding New York City (NYC). First, the trends in at-home tests that were voluntarily reported to local health departments (phone, email, online) added to the public health surveillance system relative to laboratory-based testing. Second, we aimed to understand the proportion of persons with at-home and laboratory-based tests. Third, persons who reported at-home tests only were compared to laboratory-based tests only by demographics, K-12 school-based attendance or workplace, vaccination status, severe disease, symptoms, and knowledge of where exposed (the source of infection).

## 2. Materials and Methods

### 2.1 Materials

This study is based on five linked NYS databases (14, 15). The primary database for COVID-19 case investigation in the NYS, outside NYC, is the Communicable Disease Case Management System (CDCMS). The Electronic Clinical Laboratory Reporting System (ECLRS) contains all reportable (positive and negative) COVID-19 test results (nucleic acid amplification test [NAAT] or antigen) in the NYS. Home tests have not been reported to the ECLRS. Applicable information on all positive laboratory tests is automatically transmitted from ECLRS to CDCMS, and a new case in CDCMS is created if the positive test date is greater than 90 days from the specimen collection date of the first positive result or the most recent specimen collection date (16). Additionally, Local Health Departments (LHDs) can manually enter cases with positive tests (NAAT or antigen) that are not reported via ECLRS. These could be at-home tests reported to LHDs or laboratory reports from other jurisdictions. LHDs collect positive home test results via multiple mechanisms. Some have web portals utilized by the public to enter results, some receive results from schools, and other manual mechanisms, such as emails. These were then uploaded to the CDCMS and utilized for case management.

To determine vaccine status, cases in the CDCMS were linked to a combined database containing the Citywide Immunization Registry (CIR) and the NYS Immunization Information System (NYSIIS) COVID-19 vaccination data for residents of New York City and the rest of the NYS. These databases were linked using deterministic algorithms that matched the name and date of birth (DOB). Finally, the Health Electronic Response Data System (HERDS) is a statewide, daily electronic survey of hospitals that reports patients with COVID-19 hospitalized at inpatient facilities. HERDS reported hospital admissions were linked based on initials, sex, DOB, and patient ZIP code.

The period of the analysis started when the home test entry to the CDCMS was more systematic, from November 1, 2021, and limited to cases with completed interviews conducted by case investigators. The study period ended on April 30, 2022. The primary outcome, COVID-19 case (case), was defined as a report of a new positive severe acute respiratory syndrome coronavirus 2 (SARS-CoV-2) NAAT or antigen test (including at-home tests) result greater than 90 days from the first or previous case from a positive result (16). Manually created cases were excluded if the type of test (laboratory-based or at-home) was unclear. As not all counties in the NYS investigate at-home tests in CDCMS, the analysis was further restricted to 33 counties that routinely uploaded at-home test results to CDCMS.

Trends in reported testing for three mutually exclusive case-testing groups, based on how a case was reported as testing positive, are presented. The testing groups were home-test-only, laboratory-test-only, and both home-test and laboratory-based (both tests). Cases were classified as home-test-only versus laboratory-test-only using the test type available in the CDCMS. Home test cases were linked to ECLRS to identify those with laboratory tests within 7 days (before, same day, or after) of the at-home test being further classified as both tests. Fully vaccinated patients were defined as those who were vaccinated 14 days or more after the completion of the primary series.

### 2.2 Methods

The analysis provided descriptive statistics (demographics, geographic locations, K-12 school-based setting, vaccination status, symptoms, underlying conditions, and known exposure locations) for each of the three case-testing groups. School setting was based on those who responded that, “within the 2 weeks prior to symptom onset /collection date,” they did “visit/attend” a “school/university/childcare center.” Those persons who selected “School (Prek-12)” as the type of school or summer camp they visited were classified as “K-12 School Based.” Then they were offered a list of their roles in school (student, staff, faculty, teacher, volunteer, or visitor) and this was utilized to determine the sub-categories in “K-12 School Based” settings. Exposure type was based on a question, “in the past 14 days, have you been in contact with a COVID-19 Case” and if “yes,” type of exposures selected from an offered list. Symptoms, based on a list of options offered in the CDCMS, were classified into the following categories: (a) gastrointestinal (abdominal pain, dehydration, diarrhea, vomiting, and nausea); (b) back and muscle pain; (c) cold symptoms (chills, cough, fatigue, fever, headache, runny nose, sore throat); (d) cardiac, respiratory, and rigor (chest pain, difficulty breathing, dyspnea, wheezing, rigor, seizure), and (f) smell and taste. Exposure type and symptoms are not mutually exclusive categories, and a case can be classified into multiple exposure types and symptom categories given that a person can have multiple exposures and/or symptoms. Severe disease was measured as hospitalizations (reported to HERDS) within 14 days of the specimen collection date (that within 7 days was also examined).

Adjusted odds ratios (aOR) using multivariate logistic regression and 95% confidence intervals (CIs) for aORs were estimated for the following: (a) trends adjusted for the month, county, and age category for home-test-only compared to laboratory-test-only and home-test-only compared to both tests; and (b) home-test-only vs. laboratory-test-only further adjusted for race/ethnicity, setting, underlying conditions, pregnancy, exposure source, and symptoms. County was used to adjust for geographic differences, including home test usage, as county practices varied. Adjusting for the reported month accounts for temporal variation in the home test volume, especially with changing variants. Broad age categories (0-4 years, 5-11 years, 12-17 years, 18– 49, and 50-64 years, with over 64 years as the reference) were used. The reference category for race/ethnicity was white, and that for sex was female. The K-12 school is based on students, staff, faculty, and volunteers at K-12 schools, with the reference category of those not belonging to these groups (non-school-based). All analyses were conducted using the Statistical Analysis Software (SAS 9.4). The New York State Department of Health Institutional Review Board (IRB) determined that this surveillance activity was necessary for public health work and, therefore, did not require IRB review.

## 3. Results

### 3.1 Trend in Home-Tests

From November 2021 to April 2022, there were 592,227 confirmed COVID-19 cases with complete interviews from the 33 LHDs that actively investigated at-home tests in the CDCMS. Of these, 71,699 (12%) were from home-test-only, 515,001 (87%) were from laboratory-tests-only and 5,527 (1.0%) had both tests (**Table 1**). The percentage of home-test-only cases has increased (**Table 1, Figure 1**) from 0.7% in November 2021 to 21.6% in April 2022. As the percentage of home-test-only tests increased, those with both tests decreased from 12.5% in November 2022 to 4.3% in April 2022. School-aged children (aged 5-17 years) had a higher percentage of home test-only responses (**Figure 2**). Of those who underwent both tests, 28.1% underwent a laboratory-based test on the same day, and the majority within 7 days after the home test (67.6%).

**Table 1:**
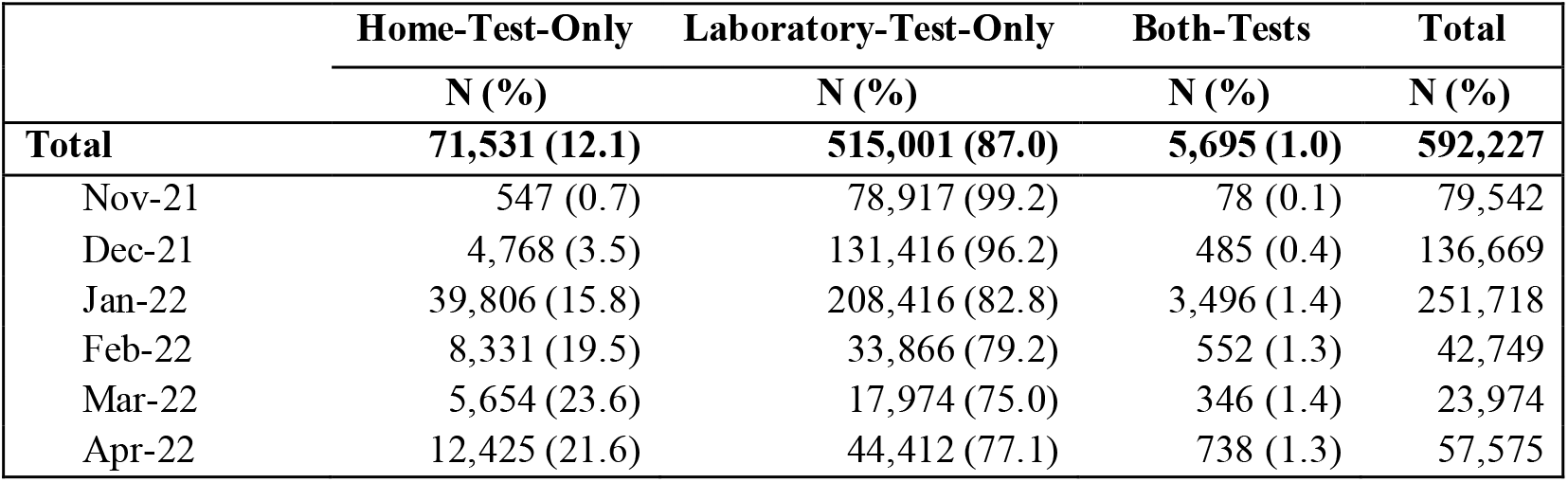
Trend in Test Type: November 2021-April 2022.

|  | **Home-Test-Only** | **Laboratory-Test-Only** | **Both-Tests** | **Total** |
| --- | --- | --- | --- | --- |
|  | **N (%)** | **N (%)** | **N (%)** | **N (%)** |
| **Total** | **71,531 (12.1)** | **515,001 (87.0)** | **5,695 (1.0)** | **592,227** |
| Nov-21 | 547 (0.7) | 78,917 (99.2) | 78 (0.1) | 79,542 |
| Dec-21 | 4,768 (3.5) | 131,416 (96.2) | 485 (0.4) | 136,669 |
| Jan-22 | 39,806 (15.8) | 208,416 (82.8) | 3,496 (1.4) | 251,718 |
| Feb-22 | 8,331 (19.5) | 33,866 (79.2) | 552 (1.3) | 42,749 |
| Mar-22 | 5,654 (23.6) | 17,974 (75.0) | 346 (1.4) | 23,974 |
| Apr-22 | 12,425 (21.6) | 44,412 (77.1) | 738 (1.3) | 57,575 |

**Figure 1:**
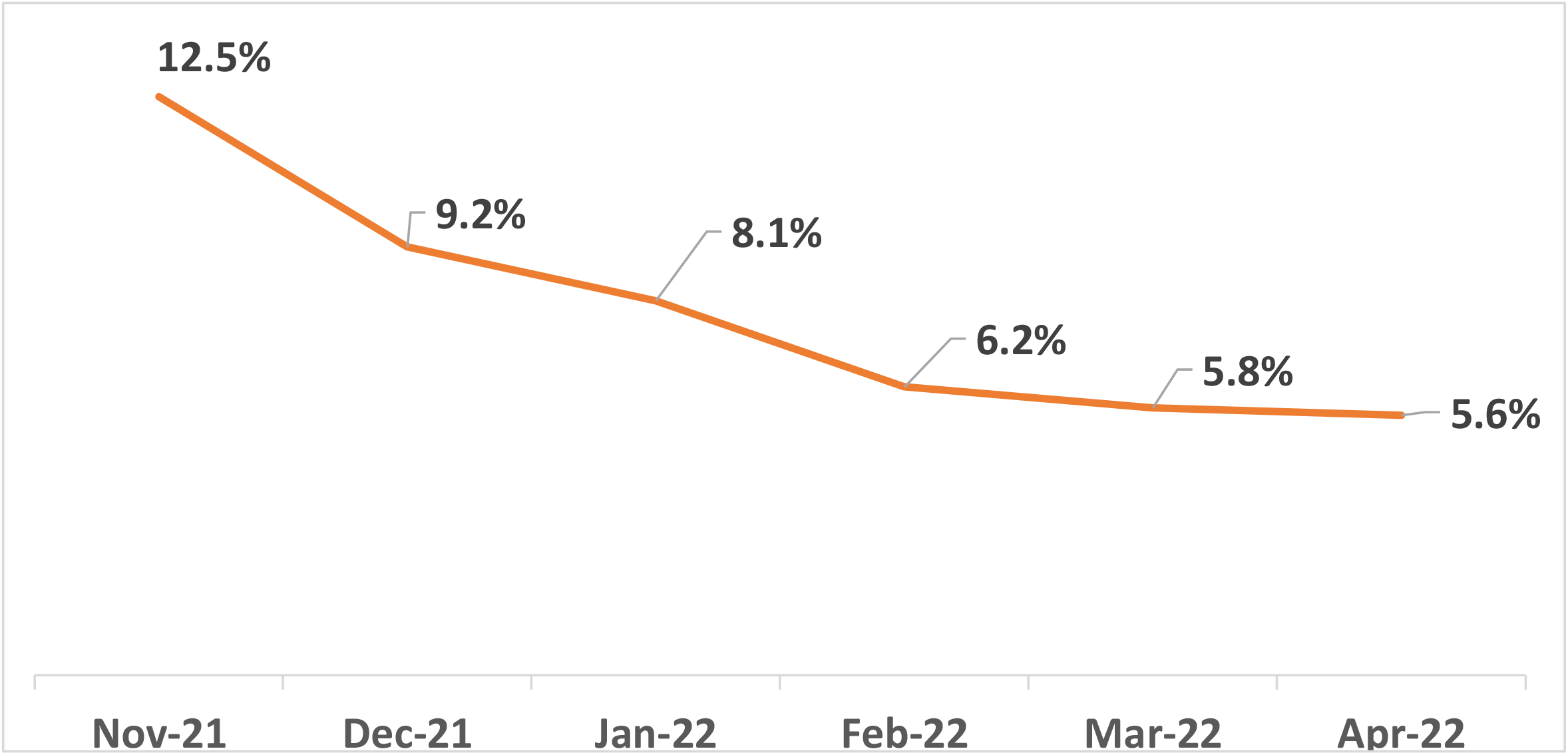
Trend in Both-Tests (Home-tests with a Laboratory Confirmed Test) as % of Any with Home Tests (Home-Tests-Only and Both-Tests): November 2021-April 2022.

**Figure 2:**
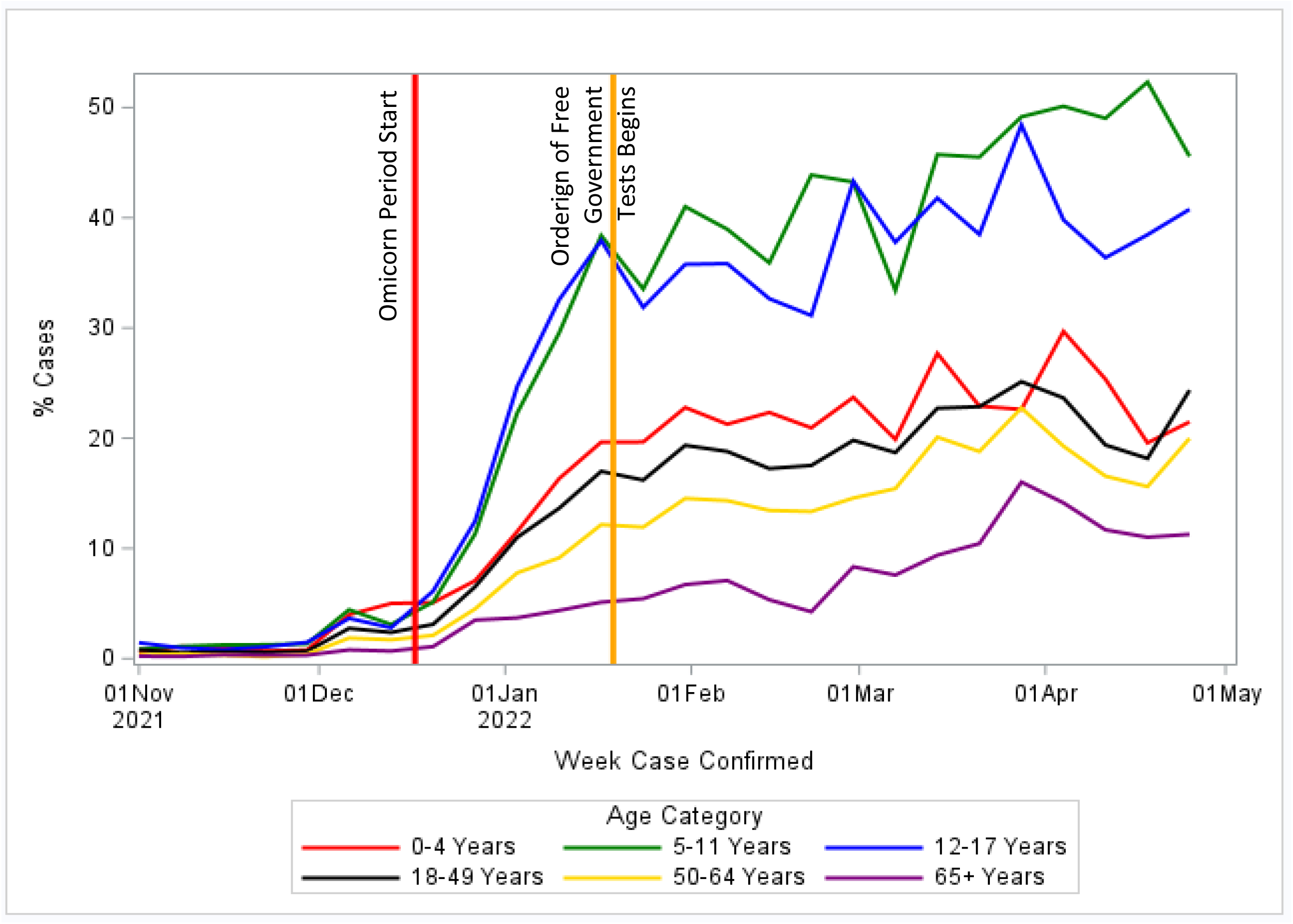
Percent via Home-Tests-Only, by Week and Age Category, November 2021-April 2022.

### 3.2 Comparison of Home-Testers vs Laboratory-Testers

Demographic characteristics are shown in **Table 2**. The percentage of unvaccinated cases was higher in the laboratory-test-only group (46.5% compared to 32.0% for both-tests and 38.7% for home-test-only), while cases with a booster dose made up a higher percentage of the at-home test group (23.6% and 26.4%, respectively, for home-test-only and both-tests compared to 15.3% for laboratory-test-only). Cases reporting working, attending, or volunteering at a K-12 school were highest among at-home tests only (41.8% home-tests-only and 26.5% for both tests compared to 19.8% for laboratory tests only). Gastrointestinal (13.6% vs. 14.7%), cardiac/respiratory/rigor (7.7% vs. 11.1%), smell/taste (6.3% vs. 13.0%) symptoms, and underlying conditions (15.0% vs. 21.4%) were lower in home-test-only cases than in laboratory-test-only cases.

**Table 2:** Profile of Laboratory-Test-Only, Home-Test-Only and Both-Tests Cases.

|  | Home-Test-Only | Lab-Test-Only | Both-Tests |
| --- | --- | --- | --- |
| Total | 71,531 | 515,001 | 5,695 |
|  | N (%) | N (%) | N (%) |
| **Age Group** |  |  |  |
| 0-4 Years | 3,854 (5.4) | 24,810 (4.8) | 158 (2.8) |
| 5-11 Years | 12,861 (18.0) | 45,863 (8.9) | 512 (9.0) |
| 12-17 Years | 11,668 (16.3) | 40,619 (7.9) | 565 (9.9) |
| 18-49 Years | 31,172 (43.6) | 252,551 (49.0) | 3,363 (59.1) |
| 50-64 Years | 9,118 (12.7) | 97,796 (19.0) | 893 (15.7) |
| 65+ Years | 2,858 (4.0) | 53,362 (10.4) | 204 (3.6) |
| **Gender** |  |  |  |
| Female | 37,175 (52.0) | 275,142 (53.4) | 3,039 (53.4) |
| Male | 29,302 (41.0) | 236,690 (46.0) | 2,193 (38.5) |
| Non-Binary | 95 (0.1) | 93 (0.0) | 8 (0.1) |
| Other | 61 (0.1) | 715 (0.1) | 6 (0.1) |
| Missing | 4,898 (6.8) | 2,361 (0.5) | 449 (7.9) |
| **Race/Ethnicity** |  |  |  |
| Hispanic | 2,741 (3.8) | 38,266 (7.4) | 230 (4.0) |
| Asian | 565 (0.8) | 9,021 (1.8) | 68 (1.2) |
| Black | 2,421 (3.4) | 28,468 (5.5) | 266 (4.7) |
| White | 45,525 (63.6) | 303,682 (59.0) | 3,385 (59.4) |
| Native American | 463 (0.6) | 2,163 (0.4) | 38 (0.7) |
| Pacific Islander | 52 (0.1) | 413 (0.1) | 6 (0.1) |
| Other | 2,101 (2.9) | 21,998 (4.3) | 141 (2.5) |
| Missing | 20,404 (28.5) | 149,256 (29.0) | 1,791 (31.4) |
| **Vaccination Status** |  |  |  |
| Unvaccinated | 27,706 (38.7) | 239,389 (46.5) | 1,821 (32.0) |
| Partial | 2,349 (3.3) | 17,059 (3.3) | 197 (3.5) |
| Primary Series Only | 24,626 (34.4) | 179,564 (34.9) | 2,173 (38.2) |
| Boosted | 16,850 (23.6) | 78,989 (15.3) | 1,504 (26.4) |
| **K-12 School Vs Non-School** |  |  |  |
| Non-School | 41,655 (58.2) | 413,205 (80.2) | 4,183 (73.5) |
| School (K-12) | 29,876 (41.8) | 101,796 (19.8) | 1,512 (26.5) |
| Faculty/Teacher | 4,437 (6.2) | 13,080 (2.5) | 342 (6.0) |
| Student | 22,442 (31.4) | 77,308 (15.0) | 929 (16.3) |
| Staff (non-Teacher) | 2,799 (3.9) | 10,339 (2.0) | 227 (4.0) |
| Volunteer/Visitor | 198 (0.3) | 1,069 (0.2) | 14 (0.2) |
| **Hospitalization** |  |  |  |
| Within 7 Days | 5 (0.0) | 13,428 (2.6) | 60 (1.1) |
| Within 14 Days | 44 (0.1) | 15,063 (2.9) | 79 (1.4) |
| **Symptoms** |  |  |  |
| Any Symptoms | 62,661 (87.6) | 418,744 (81.3) | 4,471 (78.5) |
| Total | 71,531 | 515,001 | 5,695 |
|  | N (%) | N (%) | N (%) |
| Gastrointestinal | 9,737 (13.6) | 75,775 (14.7) | 826 (14.5) |
| Back and muscle pain | 17,942 (25.1) | 133,072 (25.8) | 1,560 (27.4) |
| Cold symptoms | 55,661 (77.8) | 353,745 (68.7) | 4,031 (70.8) |
| Cardiac, respiratory and<br>rigor | 5,527 (7.7) | 56,997 (11.1) | 591 (10.4) |
| Smell and taste | 4,511 (6.3) | 66,991 (13.0) | 466 (8.2) |
| **Underlying Conditions** | 10,707 (15) | 110,332 (21.4) | 983 (17.3) |
| **Pregnant** | 304 (0.4) | 3,324 (0.6) | 46 (0.8) |
| **Exposure Source Known** | 7,868 (11.0) | 90,742 (17.6) | 605 (10.6) |
| **Known Exposure Type** |  |  |  |
| Congregate Housing | 18 (0.0) | 385 (0.1) | 2 (0.0) |
| Day Care/School | 741 (1.0) | 4,854 (0.9) | 62 (1.1) |
| Place of Employment | 499 (0.7) | 8,569 (1.7) | 65 (1.1) |
| Healthcare Facility | 21 (0.0) | 607 (0.1) | 7 (0.1) |
| Living in Same Household | 4,562 (6.4) | 46,836 (9.1) | 284 (5.0) |
| At Home, from Visitor to<br>Home | 442 (0.6) | 6,576 (1.3) | 45 (0.8) |
| Long Term Care Facility | 15 (0.0) | 479 (0.1) | 3 (0.1) |
| Political Rally/Gathering | 3 (0.0) | 18 (0.0) | - (0.0) |
| Religious Gathering | 19 (0.0) | 216 (0.0) | - (0.0) |
| Sports Event | 53 (0.1) | 312 (0.1) | 2 (0.0) |
| Social Event | 240 (0.3) | 4,672 (0.9) | 25 (0.4) |
| Travel | 21 (0.0) | 325 (0.1) | 2 (0.0) |
| Summer Camp | 0 (0.0) | 5 (0.0) | 0 (0.0) |
| Other | 505 (0.7) | 7,237 (1.4) | 64 (1.1) |
| Unknown | 82 (0.1) | 1,265 (0.2) | 8 (0.1) |

### 3.3: Adjusted Trends: Home-Test-Only vs Laboratory-Test-Only

**Figure 3** shows the aOR and CI trends adjusted for age and county for home-tests-only versus laboratory-tests-only and home-tests-only versus both tests. November was combined with December given the small number of cases in November 2021, and the figure provides the monthly trend in reference to November and December 2021. The increasing trend of home tests only was consistent. The odds ratio of home-test-only vs laboratory-tests-only goes from 8.11 (95% CI:7.86, 8.35) in January 2022 to 14.71 (95% CI:14.19, 15.26) in April 2022 relative to November/December 2021. There was a concurrent increase in home-tests-only relative to both tests (a person with an at-home test having a laboratory-based test within 7 days of the at-home test) increasing from 1.36 (95% CI:1.23, 1.31) in January 2022 to 2.45 (95% CI:2.17, 2.78) in April 2022 relative to November/December 2021.

**Figure 3:**
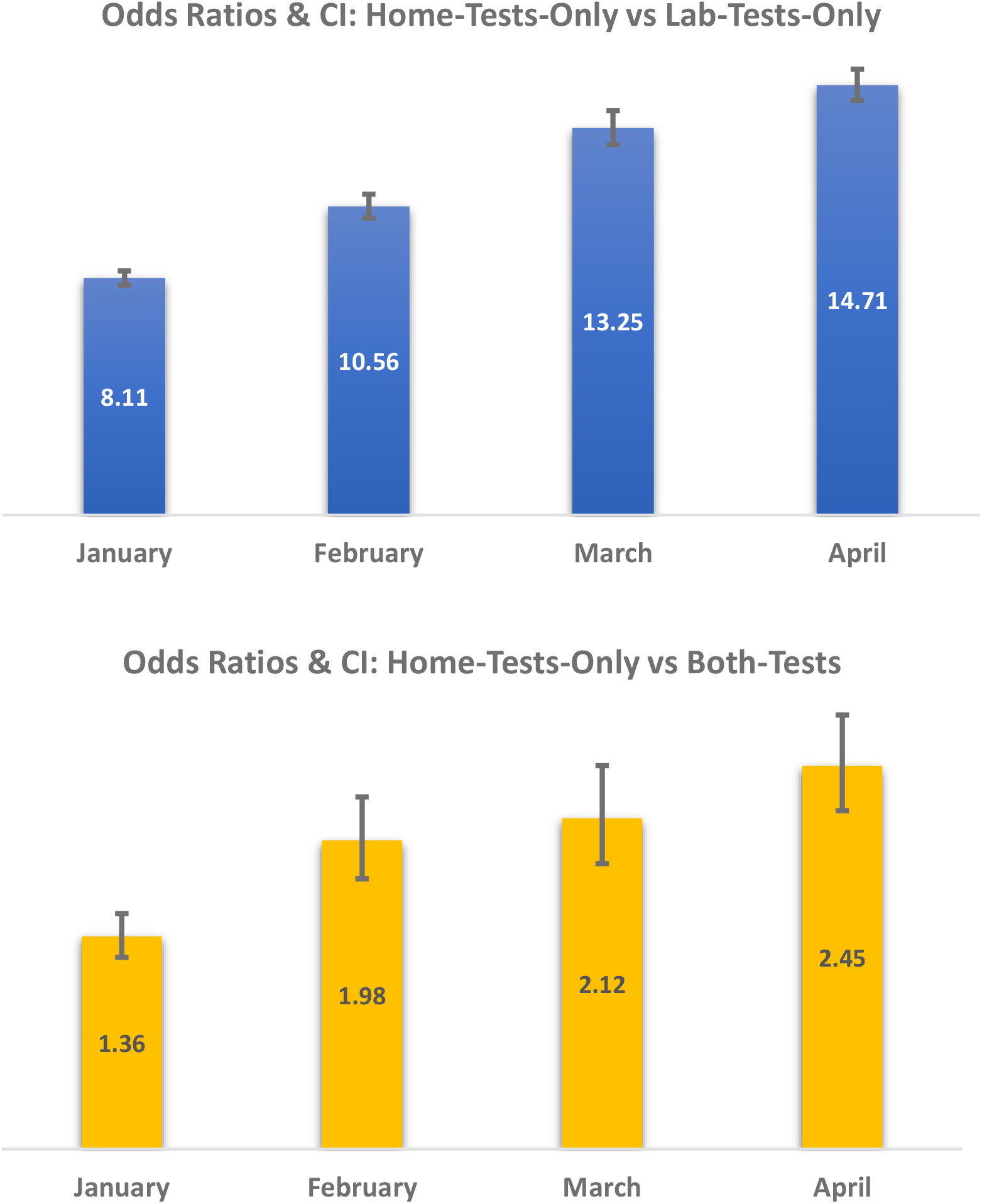
Trend Test Type for COVID-19 Cases Relative to November/December 2021 Controlled for Age and County^a^. ^a^ This is adjusted for age and county. Combined November/December 2021 were used as reference category given the small number of home-tests in November 2021 as demonstrated in table 1 and figure 2.

### 3.4: Adjusted Comparison of Home-Test-Only vs Laboratory-Test-Only

**Figure 4** demonstrates that once adjusted for the month, county, and other variables displayed, differences between home-test-only vs. laboratory-test-only (observed with descriptive statistics in **Table 2**) persisted and were statistically significant. Boosted persons were most likely to report home tests only (aOR 1.87, 95% CI:1.82, 1.93) compared to laboratory tests only, followed by those with only a primary series (aOR 1.30, 95% CI:1.27, 1.33) with unvaccinated as the reference category. The results in the partially vaccinated group were not statistically significant.

**Figure 4:**
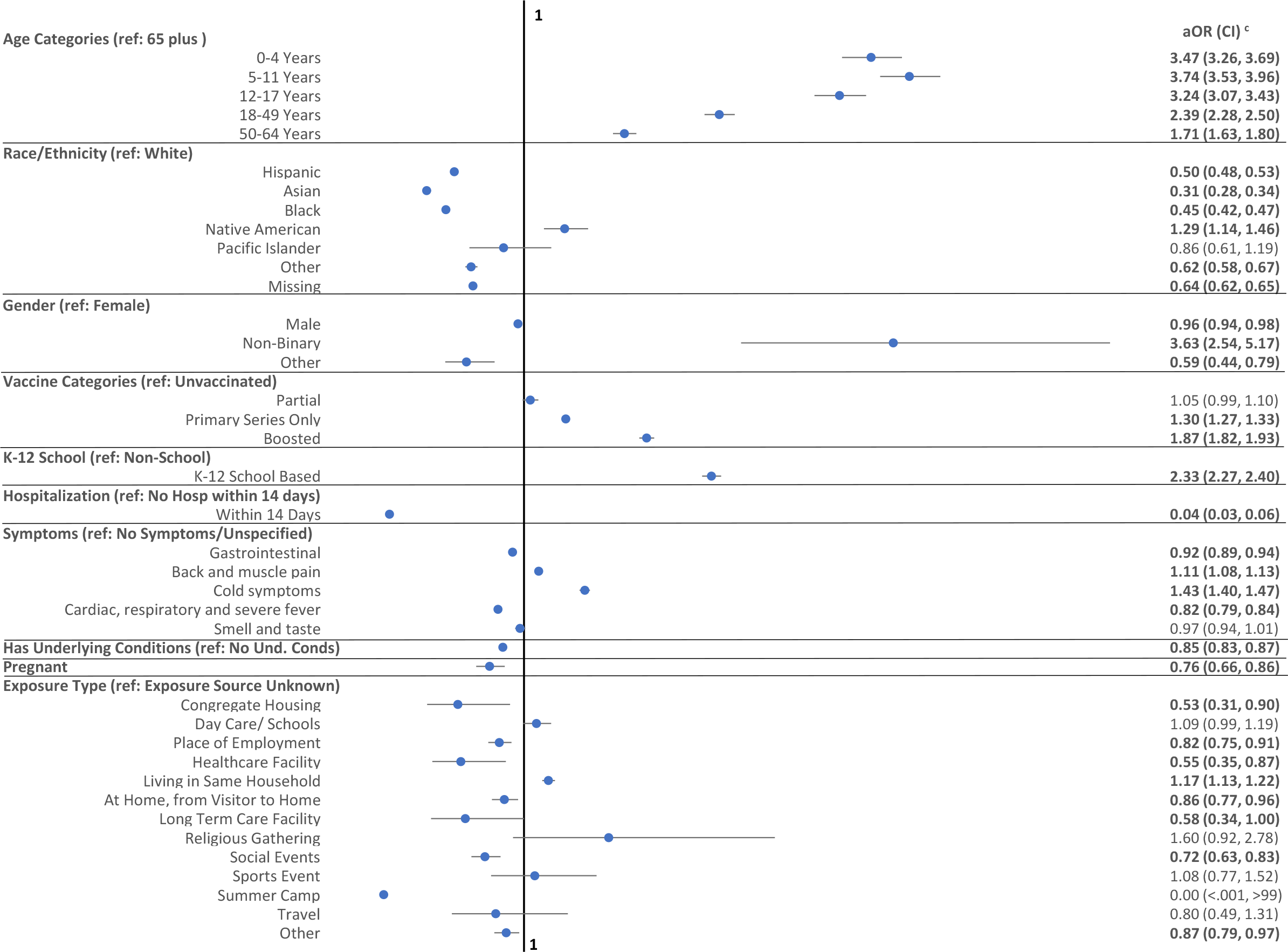
Multivariate Logistic Regression Adjusted Odds Ratios (aOR) and Confidence Intervals (CI) for Home-Tests-Only vs Laboratory Test-Only ^b^. ^b^ The model adjusted for month, county and gender missing as well as known exposure sources of political rally/gathering, summer camp and not reported, in addition to displayed variables. ^c^ Highlighted odds ratios indicate p<0.05.

Those in K-12 settings were more likely to use home tests only, whereas those in non-K-12 settings were more likely to use laboratory tests only. The odds ratio for a home-test-only was 2.33 (95% CI:2.27, 2.40) relative to laboratory-test-only for those in K-12 settings. When restricted to the school-age (5-17 years) population, the odds ratio declines to 1.55 (95% CI:95% CI:1.46, 1.65) (**Table A.2**). In contrast, when restricted to adults (>18 years) only, the odds ratio increases to 2.85 (95% CI:2.75, 2.95) (**Table A.3**).

Laboratory tests only capture severe disease as measured by hospitalization, certain symptoms, and cases with underlying conditions. Hospitalization within 14 days was less likely among home-test-only cases than laboratory-test-only cases, with only 44 home-test-only cases (aOR 0.04, 95% CI:0.03, 0.06) being hospitalized. The results for hospitalizations within seven days, with only five home-test-only cases with hospitalization, were consistent. Persons with underlying conditions (aOR 0.85, 95% CI:0.83, 0.87) or who were pregnant (aOR 0.76, 95% CI:0.66, 0.86) were less likely to have home-test-only, as were those with gastrointestinal (aOR 0.92, 95% CI:0.89, 0.94) and cardiac, respiratory, and rigor (aOR 0.82, 95% CI:0.79, 0.84) symptoms. Those with certain symptom groups (controlled for other symptoms) were more likely to have a home-test-only with 1.11 (95% CI:1.08, 1.13) for back and muscle pain and 1.43 (95% CI:1.40, 1.47) for cold symptoms relative to those without any symptoms reported.

For cases with a known exposure, the source of the exposure was associated with the testing group. If known exposure was from a household member (aOR: 1.17, 95% CI:1.13, 1.22), then the home-test-only was more likely. In contrast, those with a known exposure to a case in congregate housing (aOR 0.53, 95% CI:0.31, 0.90), place of employment (aOR 0.82, 95% CI:0.75, 0.91), a healthcare facility (aOR 0.55, 95% CI:0.35, 0.87), long term care facility (aOR 0.58, 95% CI:0.34, 1.00), or a social event (aOR 0.72, 95% CI:0.63, 0.83) were less likely to be home-test-only.

## 4. Discussion

By linking several databases, this study provides valuable insight into trends in home testing and persons excluded from official laboratory-confirmed test-based metrics in New York State (excluding New York City). The strengths of this study include the analysis of tests during the Omicron period with widespread home testing coverage and population-based large sample sizes using case investigation data collected by trained interviewers, as opposed to survey data impacted by response bias. However, testing trends as well as demographic (age, race) and vaccination status observed were consistent with survey-based studies (9, 12, 13, 17) and were able to validate each other. This study demonstrated that certain populations are more likely to use home tests only. Specifically, those at lower risk of severe disease, such as boosted, with certain symptom categories, younger age groups, persons not hospitalized, and those without underlying conditions or special medical needs, are more likely to use home tests only.

Racial differences in-home testing with a higher likelihood for Whites and a lower likelihood for Blacks, Asians, Native Americans, and Hispanic may not only reflect testing behavior, but also accessibility, knowledge, and economic differences in testing (6, 18), particularly given the fact that COVID-19 has disproportionately impacted communities of color (19-24). While data indicate that non-binary persons are more likely to utilize only home testing, it is difficult to be conclusive given the higher percentage of missing gender among home testers (6.9% for home-test-only, 0.5% laboratory-test-only) and the small number of persons identified as non-binary (95 home-test-only, 93 laboratory-test-only).

Age differentials in home testing were consistent with those in other studies on adults (9). The comparison of adults with school-age populations provides additional insight. The higher home-testing likelihood observed for the school-aged population is consistent with a recent study (33). This volume may be driven by a sharp rise in cases with Omicron onset. It may have continued with the widespread availability of test kits through the school, requirements by schools to have negative tests if symptomatic, and concerns about transmission risk to other household members (11, 25). When restricted to adults, the higher likelihood of home-tests-only for K-12 school-based persons indicates that it is not only age, but school setting that also impacts the test choice.

The likelihood of reported home tests based on vaccination status may be affected by four factors. First, employment testing requirements that may have existed for unvaccinated persons may have contributed (26). Second, the potential for more severe disease if unvaccinated persons are positive may result in a higher likelihood of them opting for laboratory-confirmed testing when presenting with symptoms. Third, given the reverse likelihood of boosting, they may have opted for home testing. It has been demonstrated that vaccination reduces severity (31, 32), and further analysis on vaccination, boosters, and severity is warranted. Fourth, individuals who are more conscientious about being vaccinated may be more likely to test at home and report.

Hospitalization, certain symptom groups, and underlying condition differentials between the home test-only and laboratory test-only cases indicate that cases captured via official metrics include those with severe disease and potential for severe disease. This indicates that publicly reported metrics capture a high proportion of, if not most, severe diseases and those with the potential for severe disease.

The association between the testing type and known exposure was not surprising. Healthcare facilities, congregate housing, and long-term care facilities are all congregate care settings in which laboratory-testing confirmation may be desired by employers and administrators for both staff and residents. While the risk of transmission was higher among household members (19, 12-29), the perception that a home test was sufficient may result in those exposed by a household member opting for only home tests. Both the increased availability of home tests via schools and similar perceptions may lead those exposed in daycare or schools to use only home tests.

This study has several limitations. First, the analysis was restricted to LHDs who were willing and able to accept home-test reports. Among them, home tests were voluntarily reported, while the comparison group included mandated reporting for laboratory-based cases. The conclusions drawn may be skewed by a person’s willingness to report home-based tests. One would expect those with severe forms of disease to seek care, and healthcare facilities would require laboratory confirmation and as such be included. For example, according to a recent survey, laboratory-confirmed tests had a 14.1% estimated test positivity rate for SARS-CoV-2 infection compared to 5.2% for exclusive home testers during the period from January 1 to March 16, 2022 (12). Second, negative home test results have not been reported for CDCMS. However, while negative laboratory-confirmed tests are reported for ECLRS, they are not part of this analysis. Thus, the impact would be minimal. Third, deterministic database matches may exclude matches when identifiers are not identical because of factors such as incorrect dates of birth, spelling of names, and maiden names versus married names. This analysis was limited to cases with completed interviews, and those with incomplete interviews may have differed (willingness to participate in disease surveillance). Finally, this analysis covers a period when universal COVID-19 case investigations and contact tracing were no longer recommended (34). As such, this study was based on COVID-19 case investigations conducted during the study period.

These findings indicate that those excluded from the official metrics are less likely to be hospitalized and have less potential for severe disease, as measured by age, vaccination status, and underlying conditions.

## Data Availability

All data produced in the present study are available upon reasonable request to the authors.

## Abbreviations

COVID 19: Coronavirus 2019
CDCMS: Communicable Disease Case Management System
ECLRS: Electronic Clinical Laboratory Reporting System
NAAT: nucleic acid amplification test
LHD: Local Health Departments
CIR: Citywide Immunization Registry
NYSIIS: NYS Immunization Information System
HERDS: Health Electronic Response Data System
SARS-COV-2: severe acute respiratory syndrome coronavirus 2

## 5. Acknowledgments

We would like to acknowledge Doris Maduka, DrPH, for her contributions in creating the figures and conducting the literature search. We would like to thank Editage (www.editage.com) for English language editing.

## 6. Funding

This work was supported by the New York State Department of Health and Centers for Disease Control (CDC), Grant #15-1043-06.

## Notes

### Competing Interest Statement

The authors have declared no competing interest.

### Author Declarations

The New York State Department of Health Institutional Review Board (IRB) determined that this surveillance activity was necessary for public health work and, therefore, did not require IRB review.

