## Supplement for "Comparison of COVID-19 Home-Testers vs Laboratory-Testers in New York State (Excluding New York City), November 2021 to April 2022"

### Data Supplement

**Figure A1: Weekly Trends in Cases by Testing Category: November 2021-April 2022**

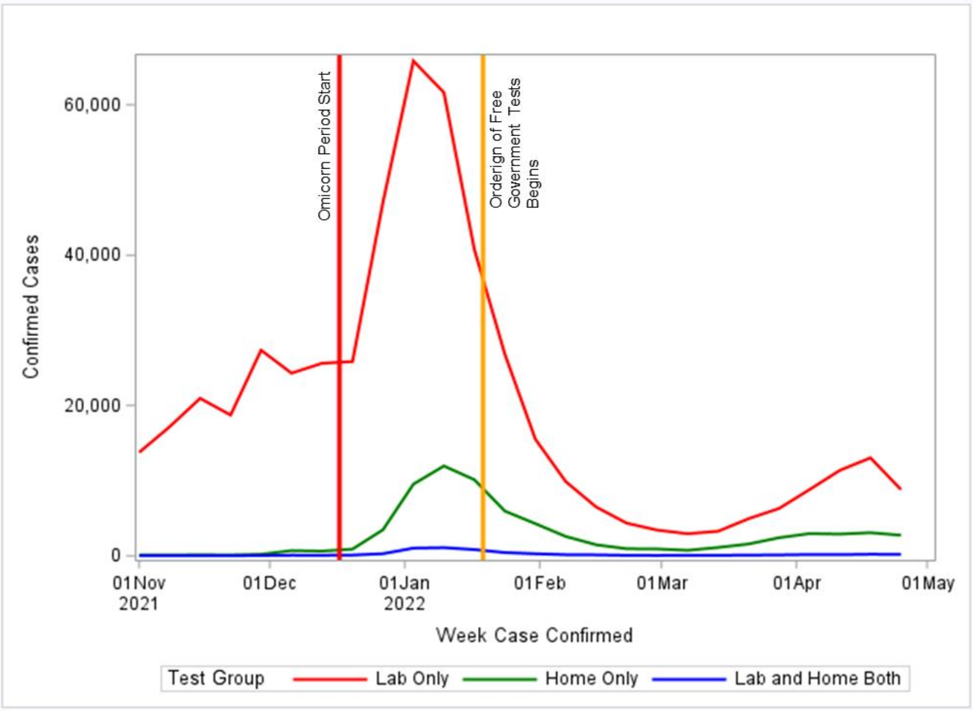


**Figure A2: Weekly Trends in % of Home-Tests of Confirmed Cases and % of Home Tests with a Laboratory-Confirmed-Test: November 2021-April 2022**

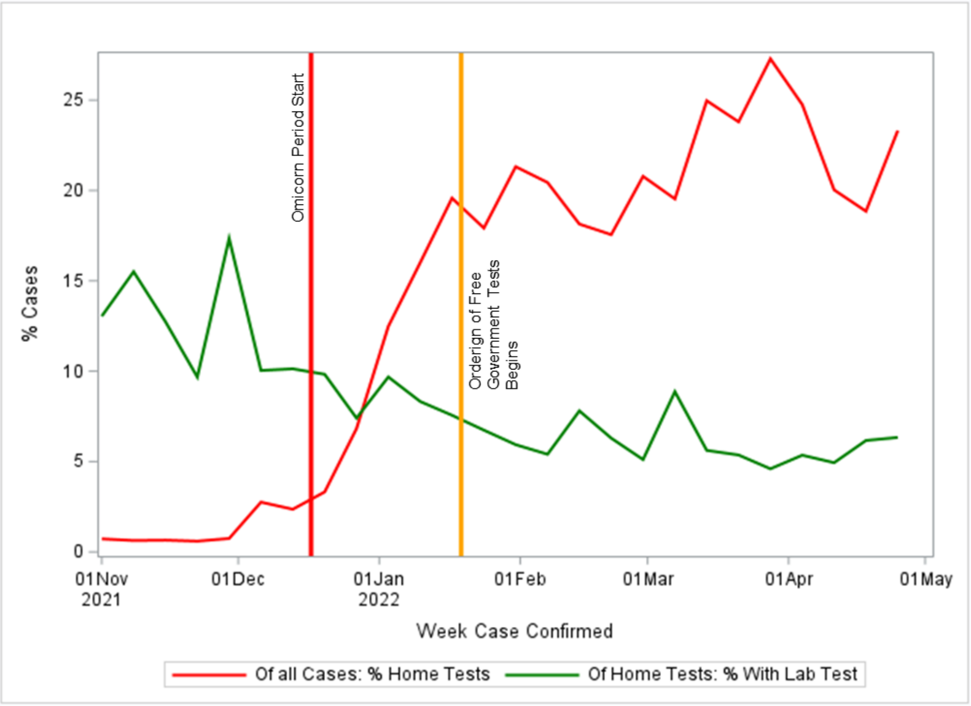

**Table A1: Laboratory Confirmed Only, Home Test Only and Both Tests Cases: Detailed Symptoms†**

| | Lab Test Only | Home Test Only | Both Tests |
|---|---|---|---|
| **Total** | **515,001** | **71,531** | **5,695** |
| | **N (%)** | **N (%)** | **N (%)** |
| Abdominal Pain | 15,329 (2.9) | 2,020 (2.8) | 146 (2.6) |
| Back Pain | 43,190 (8.4) | 4,417 (6.2) | 458 (8.0) |
| Chest Pain | 19,808 (3.8) | 1,980 (2.8) | 220 (3.9) |
| Chills | 90,869 (17.6) | 11,050 (15.4) | 1,069 (18.8) |
| Cough | 204,131 (39.6) | 29,669 (41.5) | 2,386 (41.9) |
| Dehydration | 7,993 (1.6) | 644 (0.9) | 81 (1.4) |
| Diarrhea | 36,845 (7.2) | 4,165 (5.8) | 384 (6.7) |
| Difficulty Breathing | 15,846 (3.1) | 1,221 (1.7) | 157 (2.8) |
| Fatigue | 135,820 (26.4) | 20,527 (28.7) | 1,610 (28.3) |
| Fever | 141,737 (27.5) | 20,937 (29.3) | 1,605 (28.2) |
| Headache | 168,557 (32.7) | 25,879 (36.2) | 2,094 (36.8) |
| Muscle Pain | 123,135 (23.9) | 16,900 (23.6) | 1,464 (25.7) |
| Nausea | 33,657 (6.5) | 4,331 (6.1) | 385 (6.8) |
| No Smell | 56,469 (10.9) | 3,310 (4.6) | 339 (5.9) |
| No Taste | 57,398 (11.1) | 3,845 (5.4) | 397 (6.9) |
| Rigor | 11,357 (2.2) | 1,013 (1.4) | 115 (2.0) |
| Runny Nose | 140,837 (27.3) | 22,308 (31.2) | 1,747 (30.7) |
| Seizure | 288 (0.1) | 26 (0.0) | 1 (0.0) |
| Short Breath | 29,566 (5.7) | 2,546 (3.6) | 286 (5.0) |
| Sore Throat | 133,072 (25.8) | 24,073 (33.7) | 1,923 (33.8) |
| Vomit | 19,185 (3.7) | 2,550 (3.6) | 181 (3.2) |
| Wheezing | 11,120 (2.2) | 983 (1.4) | 120 (2.1) |

**Figure A3: Detailed Symptoms: Home-Tests-Only vs Laboratory-Tests: Logistic Regression Coefficients Controlled for Age, Month and County§**

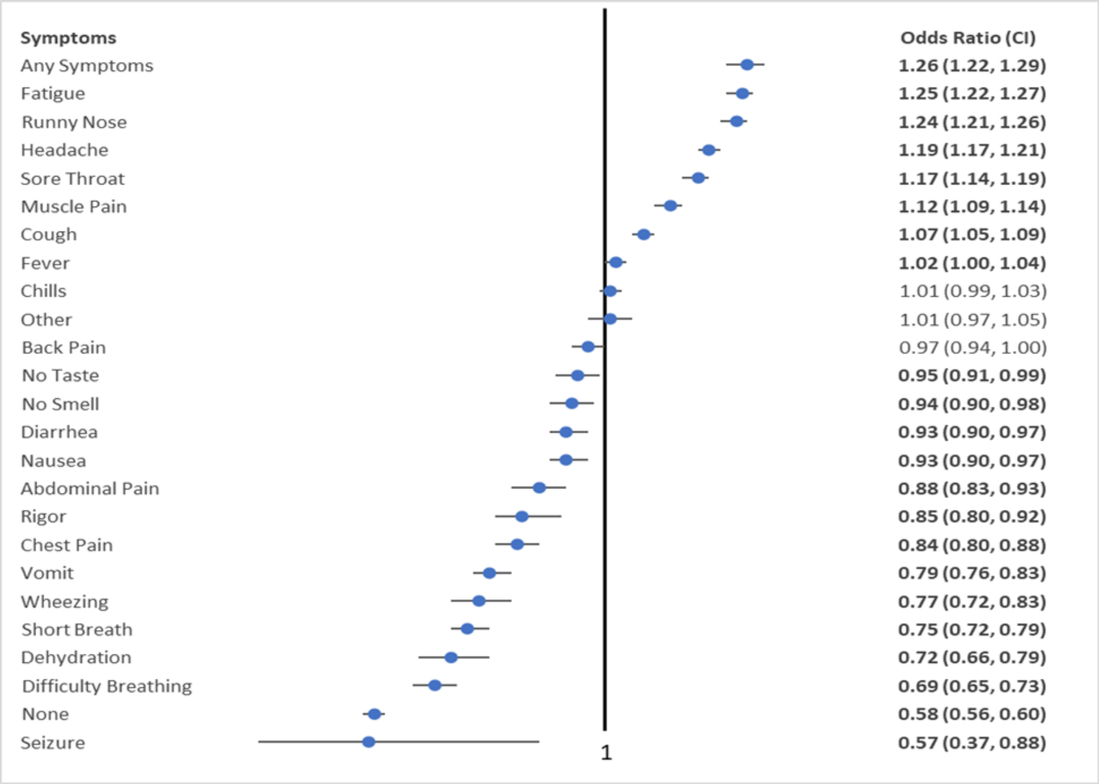


§Odd ratios were estimated separately for each symptom controlled for age, month, and county.

**Table A2: School Aged Children 5-17 Years: Home-Test-Only vs Laboratory-Test-Only, Profile and Logistic Regression Odds Ratios†**

| | Lab Test | Home Test | | |
|---|---|---|---|---|
| | N (%) | N (%) | Odds Ratio (CI) | P value |
| **Total** | **86,482 (77.9)** | **24,529 (22.1)** | | |
| **Age Group** | | | | |
| 12-17 Years | 40,619 (47.0) | 11,668 (47.6) | **Reference** | |
| 5-11 Years | 45,863 (53.0) | 12,861 (52.4) | 1.23 (1.18, 1.28) | <.0001 |
| **Vaccination Status** | | | | |
| Unvaccinated | 60,584 (70.1) | 11,769 (48.0) | **Reference** | |
| Partial | 5,221 (6.0) | 1,373 (5.6) | 1.22 (1.13, 1.31) | <.0001 |
| Primary Series Only | 19,558 (22.6) | 10,295 (42.0) | 1.65 (1.59, 1.72) | <.0001 |
| Boosted | 1,119 (1.3) | 1,092 (4.5) | 2.12 (1.91, 2.36) | <.0001 |
| **Gender** | | | | |
| Female | 42,876 (49.6) | 11,077 (45.2) | **Reference** | |
| Male | 43,153 (49.9) | 11,593 (47.3) | 1.03 (1.00, 1.07) | 0.0859 |
| Non-Binary | 24 (0.0) | 29 (0.1) | 2.81 (1.46, 5.43) | 0.002 |
| Other | 102 (0.1) | 19 (0.1) | 0.57 (0.32, 0.99) | 0.0473 |
| Missing | 327 (0.4) | 1,811 (7.4) | 47.53 (41.02, 55.08) | <.0001 |
| **Race/Ethnicity** | | | | |
| White | 45,833(53.0) | 14,501(59.1) | **Reference** | |
| Hispanic | 9,466(10.9) | 1,120(4.6) | 0.49 (0.45, 0.53) | <.0001 |
| Asian | 1,889(2.2) | 251(1.0) | 0.33 (0.28, 0.38) | <.0001 |
| Black | 5,327(6.2) | 774(3.2) | 0.44 (0.40, 0.48) | <.0001 |
| Native American | 391(0.5) | 138(0.6) | 1.01 (0.80, 1.27) | 0.9481 |
| Pacific Islander | 080(0.1) | 017(0.1) | 0.75 (0.42, 1.34) | 0.3336 |
| Other | 3,186(3.7) | 733(3.0) | 0.67 (0.61, 0.74) | <.0001 |
| Missing | 20,310(23.5) | 6,995(28.5) | 0.64 (0.61, 0.67) | <.0001 |
| **K-12 School Vs Non-School** | | | | |
| Non-School | 14,203 (16.4) | 3,386 (13.8) | **Reference** | |
| School (K-12) | 72,279 (83.6) | 21,143 (86.2) | 1.55 (1.46, 1.65) | <.0001 |
| **Hospitalization** | | | | |
| Within 14 Days | 316 (0.4) | 1 (0.0) | 0.01 (0.00, 0.09) | <.0001 |
| **Symptoms** | | | | |
| Gastrointestinal | 12,588 (14.6) | 2,981 (12.2) | 0.84 (0.80, 0.88) | <.0001 |
| Back and muscle pain | 12,677 (14.7) | 3,254 (13.3) | 1.00 (0.94, 1.05) | 0.8388 |
| Cold symptoms | 64,093 (74.1) | 18,721 (76.3) | 1.32 (1.25, 1.38) | <.0001 |
| Cardiac, respiratory and rigor | 4,058 (4.7) | 762 (3.1) | 0.73 (0.67, 0.81) | <.0001 |
| Smell and taste | 6,697 (7.7) | 795 (3.2) | 0.95 (0.86, 1.03) | 0.219 |
| **Underlying Conditions** | 10,425 (12.1) | 2,198 (9.0) | 0.85 (0.80, 0.90) | <.0001 |
| **Pregnant** | 36 (0.0) | 5 (0.0) | 0.81 (0.26, 2.53) | 0.7195 |
| **Exposure Type*** | | | | |
| Congregate Housing | 38 (0.0) | 8 (0.0) | 0.82 (0.34, 2.00) | 0.6661 |
| Day Care/School | 2,770 (3.2) | 455 (1.9) | 1.02 (0.90, 1.15) | 0.7734 |
| Place of Employment | 38 (0.0) | 6 (0.0) | 0.92 (0.34, 2.45) | 0.8605 |
| Healthcare Facility | 8 (0.0) | 4 (0.0) | 13.94 (2.94, 66.12) | 0.0009 |

| | Lab Test | Home Test | | |
|---|---|---|---|---|
| | N (%) | N (%) | Odds Ratio (CI) | P value |
| **Total** | **86,482 (77.9)** | **24,529 (22.1)** | | |
| Living in Same Household | 11,925 (13.8) | 1,874 (7.6) | 1.07 (1.01, 1.14) | 0.0337 |
| At Home, from Visitor to Home | 1,038 (1.2) | 132 (0.5) | 0.75 (0.61, 0.92) | 0.0062 |
| Long Term Care Facility | 6 (0.0) | 1 (0.0) | 0.58 (0.06, 6.00) | 0.6492 |
| Political Rally/Gathering | - | - | - | - |
| Religious Gathering | 22 (0.0) | 3 (0.0) | 0.98 (0.21, 4.53) | 0.9752 |
| Social Event | 645 (0.7) | 73 (0.3) | 0.69 (0.53, 0.91) | 0.009 |
| Sports Event | 179 (0.2) | 42 (0.2) | 1.18 (0.78, 1.77) | 0.4311 |
| Summer Camp | 4 (0.0) | - | <.001 (<.001, >99) | 0.9911 |
| Travel | 42 (0.0) | 7 (0.0) | 1.58 (0.61, 4.11) | 0.3467 |
| Other | 1,388 (1.6) | 191 (0.8) | 0.79 (0.67, 0.94) | 0.0082 |
| Unknown | 210 (0.2) | 26 (0.1) | 1.03 (0.64, 1.66) | 0.9058 |

† In addition to the displayed coefficients, the models controlled for month and county. The following coefficients are not displayed: missing gender as well as exposure sources of political rally/gathering, summer camp and unknown. Excluded exposure sources were not significant.

**Table A3: Adults 18 Years and Above: Home-Test-Only vs Laboratory-Test-Only, Profile and Logistic Regression Odds Ratios †**

| | Lab Test | Home Test | | |
|---|---|---|---|---|
| | N (%) | N (%) | Odds Ratio* (CI) | P Value |
| **Total** | **403,709 (90.3)** | **43,148 (9.7)** | | |
| **Vaccination Status** | | | | |
| Unvaccinated | 154,001 (38.1) | 12,085 (28.0) | **Reference** | |
| Partial | 11,834 (2.9) | 975 (2.3) | 0.94 (0.88, 1.02) | 0.1399 |
| Primary Series Only | 160,004 (39.6) | 14,330 (33.2) | 1.11 (1.08, 1.15) | <.0001 |
| Boosted | 77,870 (19.3) | 1,5758 (36.5) | 1.74 (1.69, 1.80) | <.0001 |
| **Age Group** | | | | |
| 65+ Years | 53,362 (13.2) | 2,858 (6.6) | **Reference** | |
| 18-49 Years | 252,551 (62.6) | 31,172 (72.2) | 2.32 (2.22, 2.43) | <.0001 |
| 50-64 Years | 97,796 (24.2) | 9,118 (21.1) | 1.67 (1.59, 1.76) | <.0001 |
| **Gender** | | | | |
| Female | 22,0368 (54.6) | 24,359 (56.5) | **Reference** | |
| Male | 180,715 (44.8) | 15,796 (36.6) | 0.94 (0.91, 0.96) | <.0001 |
| Non-Binary | 68 (0.0) | 66 (0.2) | 4.31 (2.83, 6.57) | <.0001 |
| Other | 600 (0.1) | 42 (0.1) | 0.61 (0.43, 0.86) | 0.0045 |
| Missing | 1,958 (0.5) | 2,885 (6.7) | 36.93 (34.26, 39.80) | <.0001 |
| **Race/Ethnicity** | | | | |
| White | 227,618(56.4) | 27,141(62.9) | **Reference** | |
| Hispanic | 25,784(6.4) | 1,440(3.3) | 0.55 (0.52, 0.59) | <.0001 |
| Asian | 6,495(1.6) | 277(0.6) | 0.30 (0.26, 0.34) | <.0001 |
| Black | 19,701(4.9) | 1,384(3.2) | 0.48 (0.45, 0.52) | <.0001 |
| Native American | 1,502(0.4) | 271(0.6) | 1.42 (1.22, 1.65) | <.0001 |
| Pacific Islander | 259(0.1) | 028(0.1) | 0.96 (0.62, 1.49) | 0.8525 |
| Other | 5,242(1.3) | 401(0.9) | 0.62 (0.55, 0.69) | <.0001 |
| Missing | 117,108(29.0) | 12,206(28.3) | 0.66 (0.64, 0.68) | <.0001 |
| **K-12 School Vs Non-School** | | | | |
| Non-School | 377,775 (93.6) | 35,284 (81.8) | **Reference** | |
| School (K-12) | 25,934 (6.4) | 7,864 (18.2) | 2.85 (2.75, 2.95) | <.0001 |
| **Hospitalization** | | | | |
| Within 7 Days | 12,807 (3.2) | 5 (0.0) | **Reference** | |
| Within 14 Days | 14,398 (3.6) | 43 (0.1) | 0.05 (0.04, 0.07) | <.0001 |
| **Symptoms** | | | | |
| Gastrointestinal | 59,380 (14.7) | 6,184 (14.3) | 0.97 (0.94, 1.01) | 0.1003 |
| Back and muscle pain | 119,443 (29.6) | 14,502 (33.6) | 1.10 (1.07, 1.13) | <.0001 |
| Cold symptoms | 271,200 (67.2) | 33,955 (78.7) | 1.52 (1.47, 1.57) | <.0001 |
| Cardiac, respiratory and rigor | 51,715 (12.8) | 4,668 (10.8) | 0.83 (0.80, 0.86) | <.0001 |
| Smell and taste | 60,035 (14.9) | 3,685 (8.5) | 0.95 (0.91, 0.99) | 0.0086 |
| **Underlying Conditions** | 98,414 (24.4) | 8,322 (19.3) | 0.83 (0.81, 0.86) | <.0001 |
| **Pregnant** | 3,285 (0.8) | 299 (0.7) | 0.71 (0.62, 0.82) | <.0001 |

| | Lab Test | Home Test | | |
|---|---|---|---|---|
| | N (%) | N (%) | Odds Ratio* (CI) | P Value |
| **Total** | **403,709 (90.3)** | **43,148 (9.7)** | | |
| **Exposure Type*** | | | | |
| Congregate Housing | 333 (0.1) | 9 (0.0) | 0.43 (0.20, 0.89) | 0.0226 |
| Day Care/School | 1,296 (0.3) | 182 (0.4) | 1.00 (0.84, 1.20) | 0.999 |
| Place of Employment | 8,530 (2.1) | 493 (1.1) | 0.85 (0.76, 0.94) | 0.0011 |
| Healthcare Facility | 595 (0.1) | 15 (0.0) | 0.42 (0.24, 0.71) | 0.0014 |
| Living in Same Household | 30,667 (7.6) | 2,256 (5.2) | 1.25 (1.19, 1.32) | <.0001 |
| At Home, from Visitor to Home | 5,146 (1.3) | 266 (0.6) | 0.87 (0.76, 1.00) | 0.0513 |
| Long Term Care Facility | 473 (0.1) | 14 (0.0) | 0.63 (0.36, 1.10) | 0.1059 |
| Political Rally/Gathering | 18 (0.0) | 2 (0.0) | 2.01 (0.43, 9.44) | 0.3773 |
| Religious Gathering | 188 (0.0) | 15 (0.0) | 1.63 (0.88, 3.00) | 0.1193 |
| Social Event | 3,830 (0.9) | 154 (0.4) | 0.74 (0.62, 0.88) | 0.0006 |
| Sports Event | 132 (0.0) | 11 (0.0) | 0.87 (0.44, 1.72) | 0.6949 |
| Summer Camp | 1 (0.0) | - | 0.06 (<.001, >99) | 0.9851 |
| Travel | 277 (0.1) | 14 (0.0) | 0.68 (0.37, 1.25) | 0.2155 |
| Other | 5,457 (1.4) | 281 (0.7) | 0.93 (0.81, 1.06) | 0.2702 |

† In addition to the displayed coefficients, the models controlled for month and county. The following coefficients are not displayed: missing gender as well as exposure sources of political rally/gathering, summer camp and unknown. Excluded exposure sources were not significant.
